# DeepSOCIAL: Social Distancing Monitoring and Infection Risk Assessment in COVID-19 Pandemic

**DOI:** 10.1101/2020.08.27.20183277

**Authors:** Mahdi Rezaei, Mohsen Azarmi

**Author notes:** ^1^, ^2^. Both authors contributed equally. Corresponding Author (M. Rezaei).

## Abstract

Social distancing is a recommended solution by the World Health Organisation (WHO) to minimise the spread of COVID-19 in public places. The majority of governments and national health authorities have set the 2-meter physical distancing as a mandatory safety measure in shopping centres, schools and other covered areas. In this research, we develop a generic Deep Neural Network-Based model for automated people detection, tracking, and inter-people distances estimation in the crowd, using common CCTV security cameras. The proposed model includes a YOLOv4-based framework and inverse perspective mapping for accurate people detection and social distancing monitoring in challenging conditions, including people occlusion, partial visibility, and lighting variations. We also provide an online risk assessment scheme by statistical analysis of the Spatio-temporal data from the moving trajectories and the rate of social distancing violations. We identify high-risk zones with the highest possibility of virus spread and infections. This may help authorities to redesign the layout of a public place or to take precaution actions to mitigate high-risk zones. The efficiency of the proposed methodology is evaluated on the Oxford Town Centre dataset, with superior performance in terms of accuracy and speed compared to three state-of-the-art methods.

## 1 Introduction

THE novel generation of the coronavirus disease (COVID-19) were reported in late December 2019 in Wuhan, China. After only a few months, the virus was hit by the global outbreak in 2020. On May 2020 The World Health Organisation (WHO) announced the situation as the pandemic^1,2^. The statistics by WHO on 26 August 2020 confirms 23.8 million infected people in 200 countries. The mortality rate of the infectious virus also shows a scary number of 815,000 people.

With the growing trend of patients, there is still no effective cure or available treatment for the virus. While scientists, healthcare organisations, and researchers are continuously working to produce appropriate medications or vaccines for the deadly virus, no definite success has been reported at the time of this research, and there is no certain treatments or recommendation to prevent or cure this new disease. Therefore, precautions are taken by the whole world to limit the spread of infection. These harsh conditions have forced the global communities to look for alternative ways to reduce the spread of the virus.

Social distancing, as shown in Figure 1(a), refers to precaution actions to prevent the proliferation of the disease, by minimising the proximity of human physical contacts in covered or crowded public places (e.g. schools, workplaces, gyms, lecture theatres, etc.) to stop the widespread accumulation of the infection risk (Figure 1(b)).

**Figure 1.**
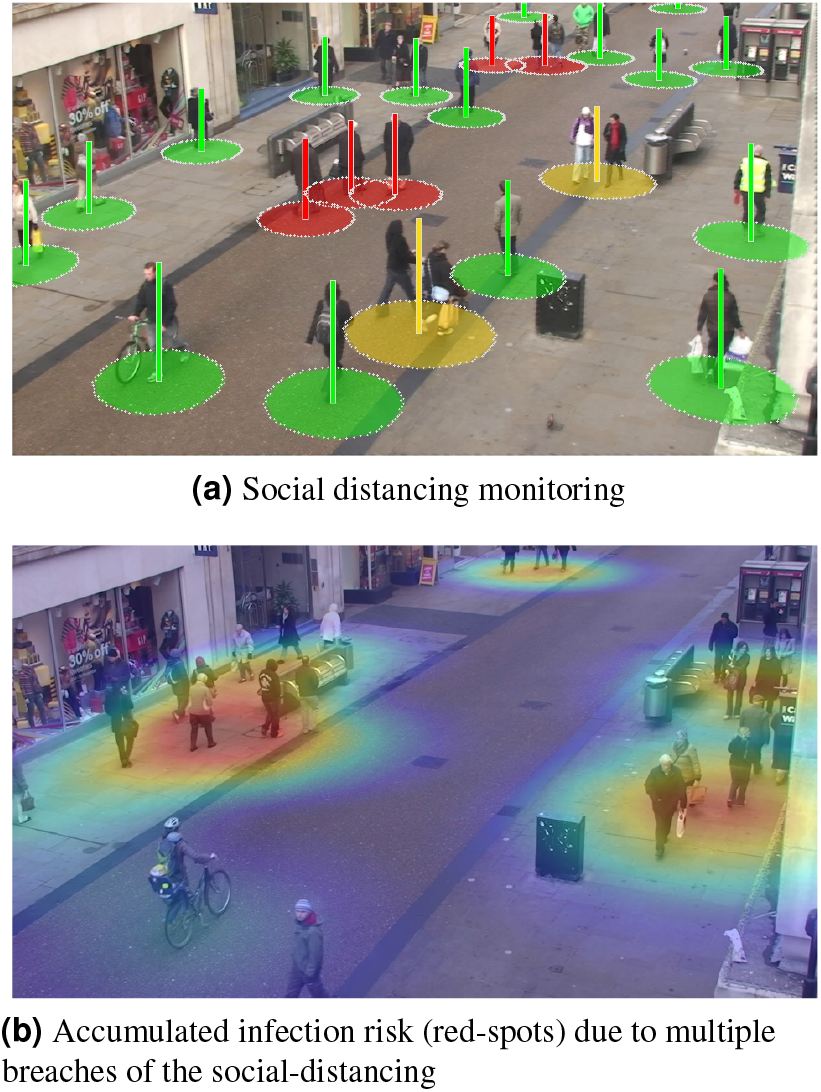
People Detection, Tracking, and Risk assessment in Oxford Town Centre, using a public CCTV camera.

According to the defined requirements by the WHO, the minimum distance between individuals must be at least 6 feet (1.8 meters) in order to observe an adequate social distancing among the people^3^.

Recent researches have confirmed that people with mild or no symptoms may also be carriers of the novel Coronavirus infections^4^. Therefore, it is important all individuals maintain controlled behaviours and observe social distancing.

Many research works such as^5,6,7^ have proved socialdistancing as an effective non-pharmacological approach and an important inhibitor for limiting the transmission of contagious diseases such as H1N1, SARS, and COVID-19.

Figure 2 demonstrates the effect of following appropriate social distancing guideline to reduce the rate of infection transmission among individuals^8,9^. A wider Gaussian curve with a shorter spike within the range of the health system service capacity makes it easier for patients to fight the virus by receiving continuous and timely support from the health care organisations. Any unexpected sharp spike and rapid infection rate (such as the red curve in Figure 2), will lead to the service failure, and consequently, exponential growth in the number of fatalities.

**Figure 2.**
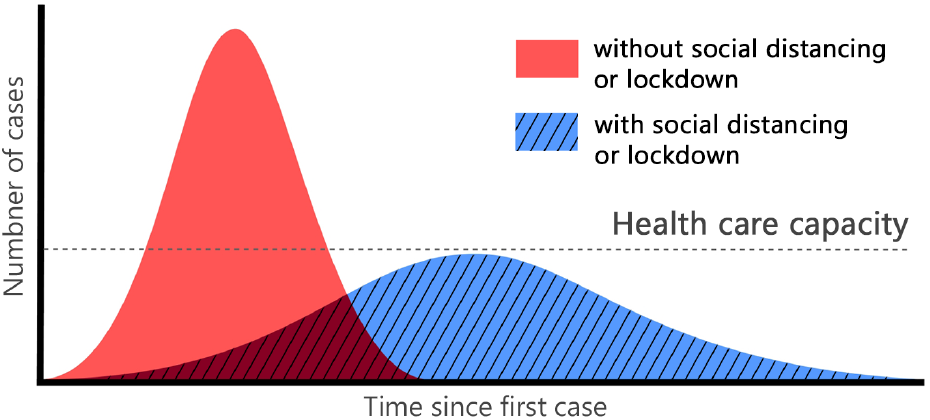
Gaussian distribution of infection transmission rate for a given population, with and without social distancing obligation.

For several months, the World Health Organisation believed that COVID-19 is only transmittable via droplets emitted when people sneeze or cough and the virus does not linger in the air. However, on 8 July 2020, the WHO announced:

> *“There has emerging evidence that COVID-19 is an airborne disease that can be spread by tiny particles suspended in the air after people talk or breathe, especially in crowded, closed environments or poorly ventilated settings.”*

Therefore, social distancing now claims to be even more important than thought before, and as one of the best ways to stop the spread of the disease in addition to wearing facial masks. Almost all countries are now considering it as a mandatory practice.

During the COVID-19 pandemic, governments have tried to implement a variety of social distancing practices, such as restricting travels, controlling borders, closing pubs and bars, and alerting the society to maintain a distance of 1.6 to 2 meters from each other^10^. However, monitoring the amount of widespread and efficiency of the constraints is not an easy task. People require to go out for essential needs such as food, health care and other necessary tasks and jobs.

In such situations, Artificial Intelligence can play an important role in facilitating social distancing monitoring. Computer vision, as a subset of Artificial Intelligence, has been very successful in solving various complex health care problems and has shown its potential in Chest CT-Scan or X-Ray based COVID-19 recognition^11,12^. Besides, deep neural networks enable us to extract complex features from the data so that we can provide a more accurate understanding of the images by analysing and classifying these features. Examples include diagnosis, clinical management and treatment, as well as the prevention and control of COVID-19^13,14^.

The main contribution of this research can be highlighted as follows:

- This study aims to support the reduction of the corona virus spread and its economic costs by providing an AI-based solution to automatically monitor and detect violations of social distancing among individuals.
- We develop one of the most (if not the most) accurate deep neural network (DNN) models for people detection, tracking, and distance estimation called *DeepSOCIAL*.
- We perform a live and dynamic risk assessment, by statistical analysis of spatio-temporal data from the people movements at the scene.
- The developed model is a generic human detection and tracker, not limited to social-distancing monitoring, and can be applied for various real-world applications such as pedestrian detection in autonomous vehicles, human action recognition, anomaly detection, and security systems.

More details and further information will be provided in the following sections. In Section 2 we discuss the related work, existing challenges, and research gaps in the field. The proposed mythology including the model architecture and our object detection techniques, tracking, and red-zone prediction algorithm will be proposed in Section 3. In Section 4 experimental results and performance of the system is investigated against the state-of-the-art, followed by discussions and concluding remarks in Section 5.

## 2 Related Works

Many researchers have worked in the medical and pharmaceutical fields aiming at treatment of COVID-19 infectious disease; however, no definite solution has yet been found. On the other hand, controlling the spread of such an unknown respiratory infectious disease is another issue.

Variety of studies with different implementation strategies^5,6,15^ have proven that controlling the prevalence is a contributing factor, and social distancing is an effective way to reduce the transmission and prevent the spread of the virus in society.

Several researchers such as^16^ and^15^ use SIR (Susceptible, Infectious, or Recovered) model. SIR is an epidemiological modelling system to compute the theoretical number of people infected with a contagious disease in a given population, over time. One of the oldest yet common SIR models is Kermack and McKendrick models introduced in 1927^17^. Eksin et al.^16^, have recently introduced a modified model of SIR by including a social distancing parameter, which can be used to determine the number of infected and recovered individuals.

Effectiveness of social distancing practices can be evaluated based on several standard approaches. One of the main criteria is based on the reproduction ratio, *Ro*, which indicates the average number of people who may be infected from an infectious person during the entire period of the infection^18^. Any *R_o_ >* 1 indicates an increasing rate of infection within the society and *R_o_ <* 1 indicates that every case will infect less than 1 person, hence, the disease rate is considered to be declining in the target population.

Since the *R_o_* value indicates the disease outspread, it is one of the most important indicators for selecting social distancing criteria. In the current COVID-19 pandemic, the World Health Organisation estimated the *R_o_* rate would be in the range of 22.5^19^, which is significantly higher than other similar diseases such as seasonal flu with *R_o_* = 1.4. In^20^, a clear conclusion is drawn about the importance of applying social distancing for the cases with a high amount of *R_o_*.

In another research based on the game theory on the classic SIR model, an assessment of the benefits and economic costs of social distancing has been examined^21^. The results also show that in the case of *R_o_ <* 1, social distancing would cause unnecessary costs, while *R_o_* ≈ 2 implies that social distancing measures have the highest economic benefits. In a similar research, Kylie et al.^22^ investigated the relationship between the stringency of social distancing and the region’s economic status. This study suggests although preventing the widespread outbreak of the virus is necessary, a moderate level of social activities could be allowed.

Prem et al.^23^ use location-specific contact patterns to investigate the effect of social distancing measures on the prevalence of COVID-19 pandemic in order to remove the persistent path of disease outbreak using susceptible-exposed-infected-removed models (SEIR).

Since the onset of coronavirus pandemic, many countries have used technology-based solutions, to inhibit the spread of the disease^24,25,26^. For example, some of developed countries, such as South Korea and India, use GPS data to monitor the movements of infected or suspected individuals to find any possible exposure among the healthy people. The India government uses the *Aarogya Setu* program to find the presence of COVID-19 patients in the adjacent region, with the help of GPS and Bluetooth. This may also help other people to maintain a safe distance from the infected person^27^. Some law enforcement agencies use drones and surveillance cameras to detect large-scale rallies and have carried out regulatory measures to disperse the population^28,29^. Some other researchers such as Xin et al.^30^ perform human detection using wireless signals by identifying phase differences and change detection in amplitude wave-forms. However this requires multiple receiving antennas and can not be easily integrated in all public places.

The utilisation of Artificial Intelligence, Computer Vision, and Machine Learning, enables us to discover the correlation of high-level features. For example, it enables us to understand and predict pedestrian behaviours in traffic scenes, sports actions and activities, medical imaging, anomaly detection, etc. by analysing Spatio-temporal visual information and statistical data analysis of the images sequences^31,14^.

In health-related fields, it would be also feasible to predict the sickness trend of specific areas^32^, to predict the density of people in public places^33^, or determine the distance of individuals from the popular swarms^34^, using a combination of visual and Geo-location cellular information. However, such research works suffer from challenges such as skilled labour or the cost of designing and implementing the infrastructures.

On the other hand, recent advances in AI, Computer Vision, Deep Learning, and Pattern Recognition, enables the computers to understand and interpret the visual data from digital images or videos. It also allows computers to identify and classify different type of objects^35,36,37^. Such capabilities can play an important role in empowering, encouraging, and performing social distancing surveillance and measurements as well. For example, computer vision could turn CCTV cameras in the current infrastructure capacity into “smart” cameras that not only monitor people but can also determine whether people follow the social distancing guidelines or not. Such systems require a very precise human detection algorithms.

People detection in image sequences is one of the most important sub-branches in the field of object detection and computer vision. Although many research works have been done in the field of human detection^38^ and human action recognition^39^, majority of them are either limited to indoor applications or suffer from accuracy issues under outdoor challenging lighting conditions. A range of other research rely on manual tuning methodologies to identify people activities, however, limited functionality has always been an issue^40^.

Convolutional Neural Networks (CNN) have played a very important role in feature extraction and complex object classification. With the development of faster CPUs, GPUs, and extended memory capacities, CNNs allow the researchers to make accurate and fast detectors compared to conventional models. However, the long time training, detection speed and achieving better accuracy, are still remaining challenges to be solved.

Narinder et al.^41^ used a deep neural network (DNN) based detector, along with Deepsort^42^ algorithm as an object tracker for people detection to assess the distance violation index. However, no statistical analysis of the outcome of their results is provided. Furthermore, no discussion about the validity of the distance measurements is provided.

In another study by Prateek et al.^43^, the authors have addressed the people distancing in a given manufactory. They have used MobileNet V2 network^44^ as a lightweight detector to reduce computational costs, which in turn provides less accuracy comparing to some other common models.

Similar to the other research, no statistical analysis is performed on the results of the distance measurement in^45^. The authors have made a comparison between two common types of DNN models (YOLO and Faster R-CNN); however, the system has been only tested on a basic dataset.

Since the topic is very recent, there has not been much research regarding the accuracy of detections, no experimental on challenging datasets has been performed, no standard comparison has been conducted on common datasets, and no analytical studies or post-processing have been considered after the detection-phase.

Considering the above-mentioned research gaps, we propose a new model which not only preforms more accurate and faster than the state-of-the-art but also will be trained and tested using large and comprehensive datasets, in challenging environments and lighting conditions. This will ensure the model is capable of performing in real-world scenarios, particularly in covered shopping centres where the lighting conditions are not as ideal as the outdoor lighting. Furthermore, we offer post-detection and post-processing analytical solutions to mitigate the spread of the virus.

## 3 Methodology

We propose a 3-stage model including people detection, tracking, inter-distance estimation as a total solution for social distancing monitoring and zone-based infection risk analysis. The system can be integrated and applied on all type of CCTV surveillance cameras with any resolution from VGA to Full-HD, with real-time performance.

### 3.1 People Detection

Figure 3 shows the overall structure of the Stage 1. The objective is to develop a model to detect humans (people) with various types of challenges such as variations in clothes, postures, at far and close distances, with/without occlusion, and under different lighting conditions.

**Figure 3.**
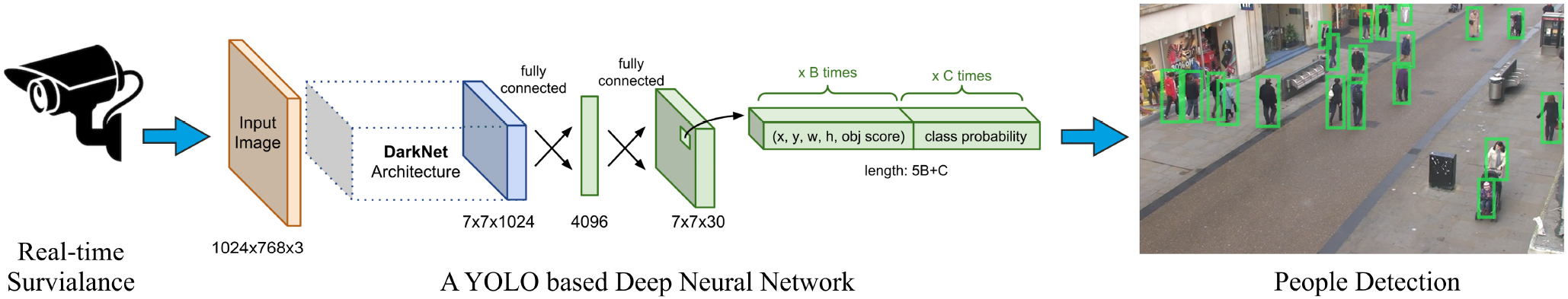
Stage 1- The overall structure of the people detection module.

To gain this we inspire from the strength of cutting-edge research; however, we develop our own unique human classifier and train our model based on a set of comprehensive and multifaceted datasets. Before diving into further technical details, we overview the most advanced object detection techniques and then introduce our human detection model.

Modern DNN-based detectors consist of two sections: A *backbone* for extracting features and a *head* for predicting classes and location of objects. The feature extractor tends to encode model inputs by representing specific features that help it to learn and discover the relevant patterns related to the query object(s). Examples of feature extraction architectures can be seen in VGG16^46^, ResNet-50^47^, CSPResNeXt-50^48^, CSPDarknet53^48^, and EfficientNet-B0/B7^49^. The head of a DNN is responsible for the classifying the objects (e.g. people, bicycles, chairs, etc.) as well as calculating the size of the objects and the coordinates of the correspondent bounding boxes.

There are usually two types of head sections: one-stage and two-stage. The two-stage detectors use the region proposal before applying the classification. First, the detector extracts a set of object proposals (candidate bounding boxes) by a selective search. Then it resizes them to a fixed size before feeding them to the CNN model. This is similar to R-CNN based detectors^50-52^. There are usually two types of head sections: one-stage and two-stage. The two-stage detectors use the region proposal before applying the classification. First, the detector extracts a set of object proposals (candidate bounding boxes) by a selective search. Then it resizes them to a fixed size before feeding them to the CNN model. This is similar to R-CNN based detectors^50-52^. In spite of the accuracy of two-stage detectors, such methods are not suitable for the systems with restricted computational resources^53^.

On the other hand, the one-stage detectors perform a single detection process such as the work done by Liu et al. (known as SSD), or other works done by Redmon et al.^54,55,56^, and Bochkovski et al^57^, known as “You Only Looks Ones” or YOLO detectors. Such detectors use regression analysis to calculate the dimensions of bounding boxes and interpret their class probabilities. It maps the image pixels to the enclosed grids and checks the probability of the existence of an object in each cell of the grids. This approach offers excellent improvements in terms of speed and efficiency.

Figure 4, shows the outcome of our investigations and reviews on some of the most successful object detection models such as RCNN^50^, fast RCNN^51^, faster RCNN^52^, SSD^58^, YOLOv1-v4^54-57^ tested on the PASCAL-VOC^59^ and MS-COCO^60^ data sets under similar conditions.

**Figure 4.**
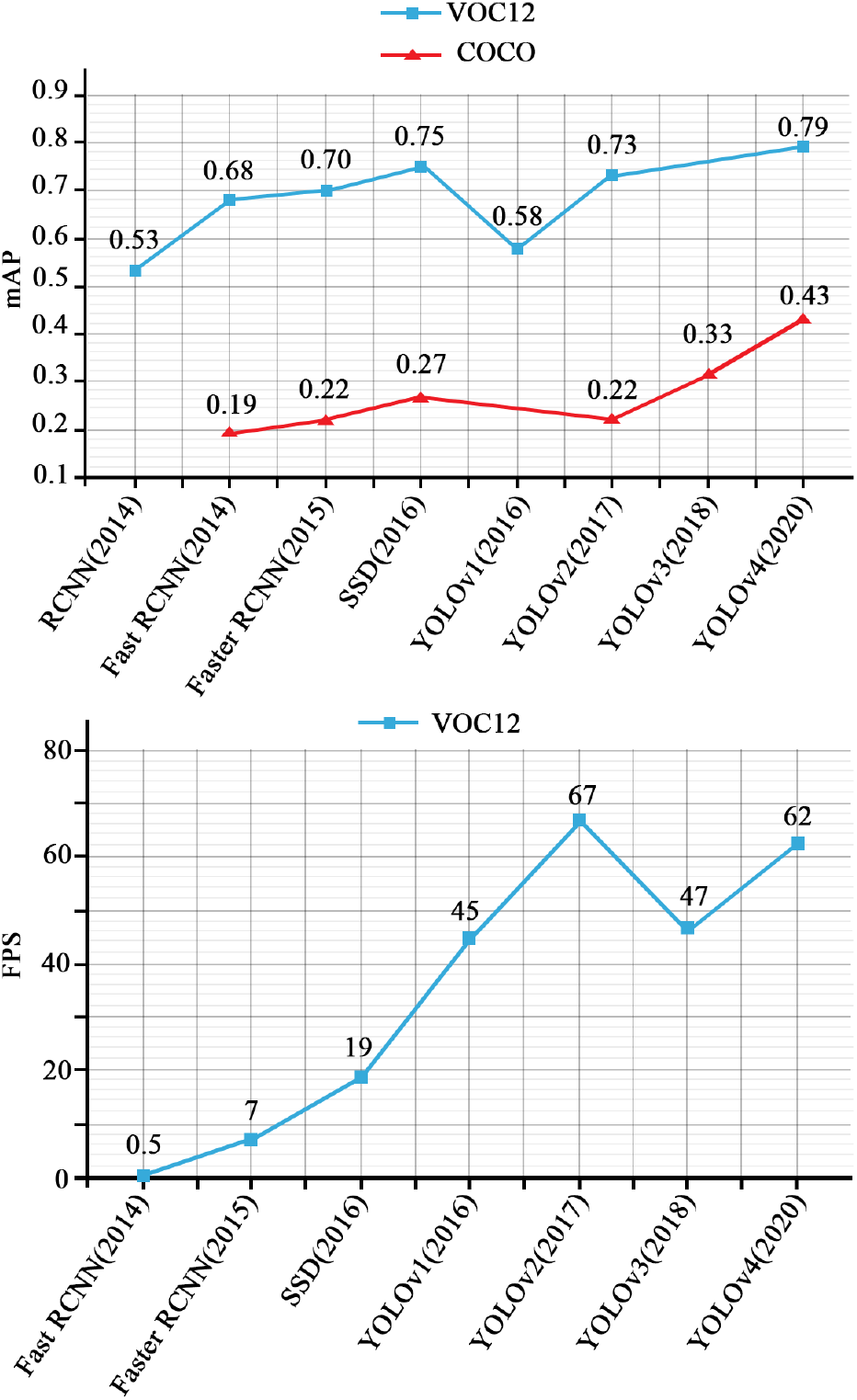
Speed and Precision overview of eight most popular object detection models on MS-COCO and PASCAL-VOC datasets.

Otherwise, the performance of the systems may vary depends on various factors such as backbone architecture, input image size, resolution, model depth, software, and hardware platform.

As illustrated in Figure 4, YOLOv4 offers the best trade-off for the speed and the accuracy; however, since YOLOv4 is an aggregation of various techniques, we undertook an in-depth study of each sub-techniques to achieve the best results for our people detection model and to outperform the state-of-the-art.

We considered two approaches to improve the accuracy of the backbone: A basic way to improve the accuracy of CNN-based detectors is to expand the receptive field and enhance the complexity of the model using more layers; however, using this technique makes it harder to train the model. We suggest using a skip-connections technique for ease of training, instead.

Various models use a similar policy to make connections between layers, such as Cross-Stage-Partial-connections (CSP)^48^ or Dense Blocks (consisting of Batch Normalisation, ReLU, Convolution, etc.) in DenseNet^61^. Such models are also been used in the design of some recent backbone architectures, such as CSPResNeXt50, CSPDarknet53^48^ and EfficientNet-B3, which are our supported architectures options for YOLOv4.

Table 1 summarises the brief report of our investigations for the above-mentioned backbone architectures in terms of number of parameters and the processing speed (in *fps*) for the same input size of 512 × 512.

**Table 1.** Comparisons of three backbone models in terms of number of parameters and speed (*fps*) using an RTX 2070 GPU.

| Backbone model | Input Resolution | Number of Parameters | Speed (*fps*) |
| --- | --- | --- | --- |
| CSPResNeXt50 | $512 \times 512$ | 20.6 M | 62 |
| CSPDarknet53 | $512 \times 512$ | 27.6 M | 66 |
| EfficientNet-B3 | $512 \times 512$ | 12.0 M | 26 |

Based on theoretical justifications in^57^ and several experiments by us we concluded that CSPDarknet53 is the most optimal backbone model for our application, in spite of higher complexity (due to more number of parameters). Here, the higher number of parameters leads to the increased capability of the model in detecting multiple objects while at the same time we can maintain real-time performance.

Recently, some of the modern proposed models have placed some extra layers between the backbone and the head, called the *neck*, which is considered for feature collection from different stages of the backbone network.

The neck section consists of several top-down and bottom-up paths to collect and combine parameters of the network in different layers, in order to provide a more accurate image features for the head section. Some of the techniques used in the neck section are the Feature Pyramid Network (FPN)^62^, Path Aggregation Network (PAN)^63^ and Spatial Attention Module (SAM)^64^.

In YOLO-v4, the authors have dealt with two categories of training options for different parts of the network: *“Bag of Freebies”*, which includes a set of methods to alter the model’s training strategy with the aim of increasing the generalisation; and *“Bag of Specials”* which includes a set of modules that can significantly improve the object detection accuracy in exchange for a small increase in training costs.

Many CNN-based models use Fully-Connected layers for classification part, and consequently, they can only accept fixed dimensions of images as input. This can lead to two types of issues: firstly, we cannot deal with low resolution images and secondly, the detection of the small objects would be difficult. These are in contradiction with our objectives where we aim to have our model applicable in any surveillance cameras with any input image sizes and resolutions.

In order to deal with the first issue we can refer to existing methodologies such as Fully Convolution Networks (FCN). These has no FC-layers and therefore can deal with images with different sizes. However, to cope with the second issue (i.e. dealing with small objects), we performed a feature pyramid technique to enhance the receptive field and extract different scales of image from the backbone, and finally, performing a multi-scale detection in the head section.

Figure 5 shows the heads that we applied at different scales of the network for object detection at different sizes. YOLOv3 uses FPN^62^ to extract features of different scales from the backbone, however in YOLOv4 based approach we use a modified version of the Spatial Pyramid Pooling layer (SPP)^65^, instead of FPN, to address the issue of various spatial dimensions as well as dealing with multi-scale detection in head section.

**Figure 5.**
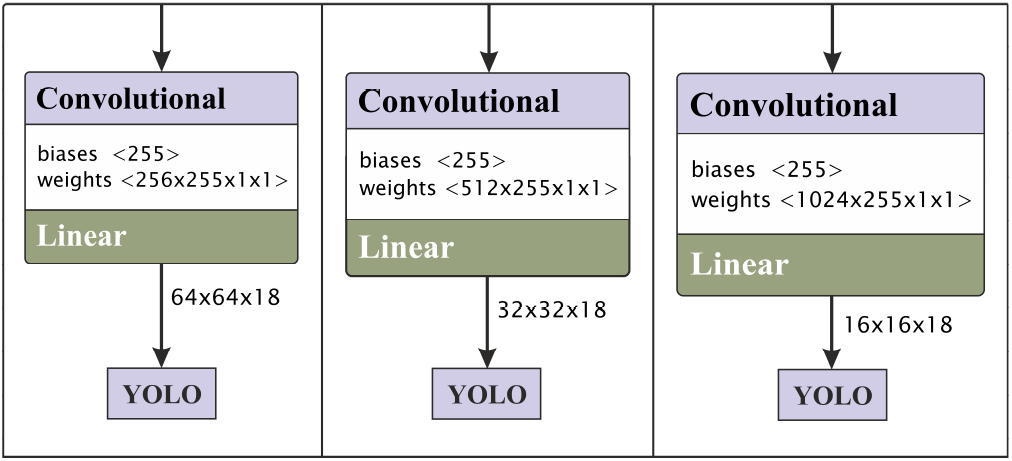
The YOLO-based heads applied at different scales

In SPP module we see the input image as a “Bag of Words” feature-map which are divided into *m* × *m* bins, where *m* can be 1, 2, or 4. A max-pooling is then applied to each bin for each channel to gets best features. In section 4 we will examine the efficiency of this approach in improving the accuracy of our model.

In DNNs, the bottom layers (i.e the first few layers) extract localised pattern and texture information to gradual build up of the semantic information which is required in the top layers. However, during the feature extraction process, some parts of the local information that may be required for fine-tuning of the model, may be lost. In PANet^63^ approach, the information flow of the bottom layers will be added to top layers to strengthen of localised information; therefore, better finetuning and prediction can be expected. In a recent research by Bochkovskiy et al.^57^, it is shown that the concatenation operator performs better than addition operator to maintain the localised information and transferring them to the top layers.

Experimenting various rational configurations for the neck module of our model, we use spatial pyramid pooling (SPP) and PAN as well as the Spatial Attention Module (SAM)^64^ which together made one of the most effective, consistent and robust components to focus the model on optimising the parameters.

In the head of the model, we use the same configuration as YOLOv3. Similar to many other anchor-based models, YOLO uses predefined boxes to detect multiple objects. Then the object detection model will be trained to predict each generated anchor boxes that belong to a particular class. After that, an offset will be used to adjust the dimensions of the anchor box in order to better match with the ground-truth data, based on the classification and regression loss.

Assuming the grid cell reference point (*c_x_,c_y_*) at the top left corner of the object image and the bounding box prior with the width and height (*p_w_, p_h_*), the network predicts a bonding box at the centre (*x, y*) and the size of (*w, h)* with the corresponding offset and scales of (*b_x_, b_y_, b_w_, b_h_*) as follows:

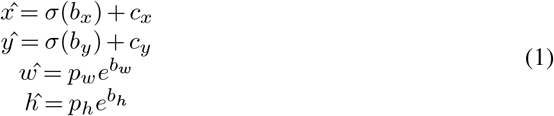

were *σ* is the Sigmoid confidence score function within the range of 0 and 1.

We represents the class of “*human*” with a 4-tuple (*x,y,w, h*), where (*x,y*) is the centre of the bounding box, and *w, h*are width and height, respectively.

We use three anchor boxes to find a maximum of three people in each grid cell. Therefore, the total number of channels is 18: (1 class + 1 object + 4 coordinates) × 3 anchors.

Since we have multiple anchor boxes for each spatial location, a single object may be associated with more than one anchor boxes. This problem can be resolved by using non-maximal suppression (NMS) technique and by computing intersection over union (IoU) to limits the anchor boxes association.

As part of the weight adjustment and loss minimisation operation, we use Complete IoU (CIoU) (in Eq.3) instead of the basic IoU (Eq. 2). The CIoU not only compares the location and distance of the candidate bounding boxes to the ground truth bounding box but also it compares the aspect ratio of the size of the generated bounding boxes with the size of the ground truth bounding box.

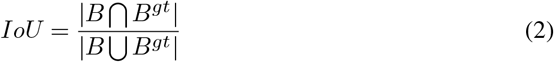

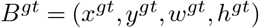 is the ground-truth box, and *B* = (*x, y, w, h*) is the predicted box. We use CIoU not only as a detection metric but also as a loss function:

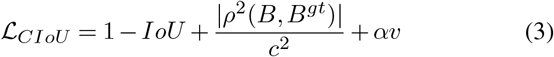

where *ρ* is Euclidean distance between grand-truth *B_gt_*, and predicted *B* bounding box. The diagonal length of the smallest bounding box enclosing both boxes *B* and *B^gt^* is represented by *c*; and *α* is a positive trade-off parameter:

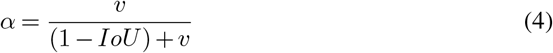

and *v* measures the consistency of the aspect ratio, as follows:

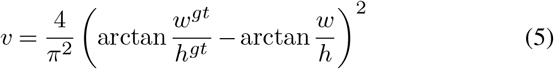

Even in case of zero-percent overlapping, the loss functions still gives us an indication on how to adjust the weights to firstly converge the aspect size towards 1, and secondly, how to reduce the error distance of candidate bounding boxes to the centre of the ground truth bounding box. A similar approach called Distance-IoU is used in^66^ for another application.

We also investigated the performance of our model against three activation functions.

**ReLU:**

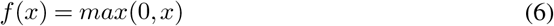

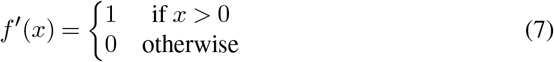

**Swish:**

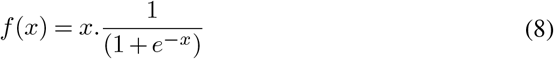

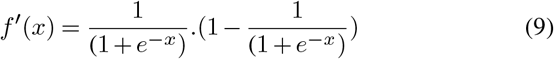

and **Mish:**

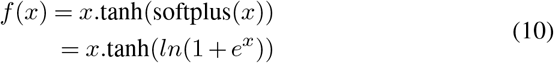

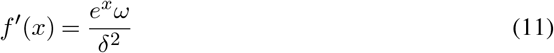

as a self regularised Non-Monotonic activation function, were

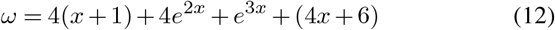

and

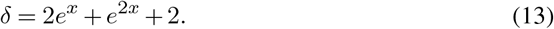

Our preliminary evaluations, confirmed the same results provided by Misra^67^ for our human detection application. The Mish activation function converged towards the minimum loss, faster than Swish and ReLU, with higher accuracy. The result was consistent especially for diversity of parameters initialisers, regularisation methods, and lower learning rate values.

Among various techniques of Bag of Freebies, we used the Mosaic data augmentations^57^ which integrates four images into one, to increase the size of the input data without requiring to increase the batch size.

On the other hand, in batch normalisation, the batch size reduction causes noisy estimation of mean and variance. To address this issue, we considered the normalised values of the previous *k* iterations instead of a single mini-batch. This is similar to Cross-Iteration Batch Normalisation (CBM)^68^.

As mentioned in^69^, using Dropblock regularisation can effectively prevent overfitting. In this regard, Class label Smoothing^70^ also helps to prevent over-fitting by reducing the model’s confidence in the training phase.

Figure 6 shows the 3-level structure of the human detection module and the sequence of the network components. In the input part, the Mosaic data augmentation (MDA), Class label smoothing (CLS) and DropBlock regularisation are applied to the input image. In the detector part, the Mish activation function has been used, and CIoU metric is considered as the loss function. In the prediction part, for each cell at each level, the anchor boxes contain the information required to locate the bounding box, confidence ratio of the object, and the corresponding class of the object. In total, we have 9 anchor boxes.

**Figure 6.**
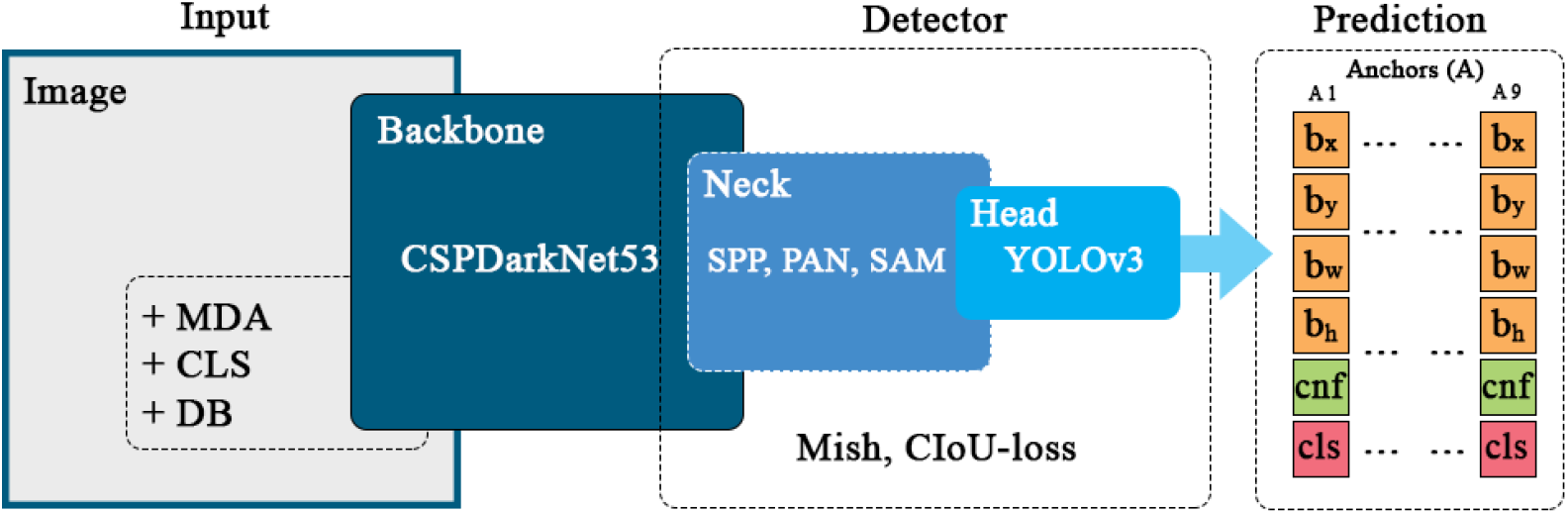
The network structure of the proposed 3-level human detection module.

We also used Stochastic Gradient Descent (SGD) with *warm restarts*^71^ to change the learning rate during the training process. This helps to jump out of local minima in the solution space and save the training time. The method initially considers a large value of the learning rate, then slows down the learning speed halfway, and eventually reduces the learning rate for each batch, with a tiny downward slope. We decreased the learning rate using a cosine annealing function for each batch as follows:

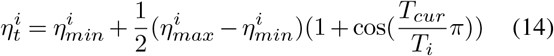

where *η_t_* is the current learning rate in the *i^th^* run, *η_min_* and *η_max_* are the minimum and maximum target learning rates. *T_cur_* is the number of epochs executed since the last restart, and *T_i_* is the number of epochs performed since the restart of the SGD.

For the training phase of the model, we considered the *person* category of the Microsoft COCO Dataset^60^ as well as the Open Images Datasets V6+^72^ which includes 16 Million ground truth bounding boxes in 600 categories. The dataset is a collection of 19,957 classes and the major part of the dataset is suitable for human detection and identification. The dataset is annotated using the bounding-box labels on each image along with the corresponding coordinates of each label.

We also considered the category of human body parts such as the legs as we believe this allows the detector to learn a more general concept of a human being, particularly in occluded situations or in case of partial visibility e.g. at the borders of the input image were the full-body of the individuals can not perceived.

### 3.2 People Tracking

The next step after the detection phase, is people tracking and ID assignment for each individual.

We use the *Simple Online and Real-time* (SORT) tracking technique^73^ as a framework for the Kalman filter^74^ along with the *Hungarian* optimisation technique to track the people. Kalman filter predicts the position of the human at time *t*+ 1 based on the current measurement at time *t* and the mathematical modelling of the human movement. This is an effective way to keep localising the human in case of occlusion.

The Hungarian algorithm is a combinatorial optimisation algorithm that helps to assign a unique ID number to identify a given object in a set of image frames, by examining whether a person in the current frame is the same detected person in the previous frames or not.

Figure 7(a) shows a sample of the people detection and ID assignment, Figure 7(b) illustrates the tracking path of each individual, and Figure 7(c) shows the final position and status of each individual after 100 frames of detection, tracking, and ID assignment. We later use such temporal information for analysing the level of social distancing violations and high-risk zones of the scene.

**Figure 7.**
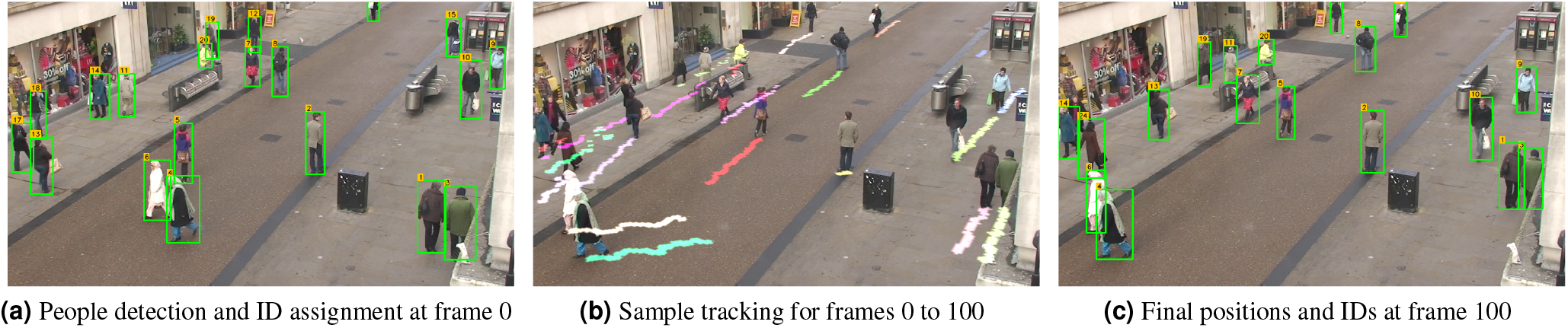
People Detection, ID assignment, Tracking and moving trajectory representation.

The state of each human in a frame is modelled as:

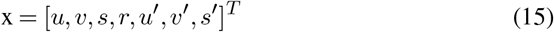

where (*u, v*) represent the horizontal and vertical position of the target bounding box (i.e. the centroid); *s* denoted the scale (area), and *r* is the aspect ratio of the bounding box sides.

When an identified human associates with a new observation, the current bounding box will be updated with the newly observed state. This will be calculated based on the velocity and acceleration components, estimated by the Kalman filter framework. If the predicted identities of the query individual significantly differ with the new observation, almost the same state that is predicted by the Kalman filter will be used with almost no correction. Otherwise, the corrections weights will be split proportionally between the Kalman filter prediction and the new observation (measurement).

As mentioned earlier, we use the Hungarian algorithm to solve data association problem, by calculating the IoU (Eq. 2) and the distance (difference) of the actual input values to the predicted values by the Kalman filter.

After the detection and tracking process, for every input frame *I_w_* _×_ *_h_* at time *t*, we define the matrix *D_t_* that includes the location of *n* detected human in the image carrier grid:

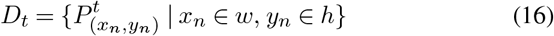

### 3.3 Inter-distance Estimation

Stereo-vision is a popular technique for distance estimation such as in^75^; however, this is not a feasible approach in our research when we aim at integration of an efficient solution, applicable in all public places using only a basic CCTV camera. Therefore we adhere to a monocular solution.

On the other hand, by using a single camera, the projection of a 3-D world scene into a 2-D perspective image plane leads to unrealistic pixel-distances between the objects. This is called perspective effect, in which we can not perceive uniform distribution of distances in the entire image. For example, parallel lines intersect at the horizon and farther people to the camera seem much shorter than the people who are closer to the camera coordinate centre.

In three-dimensional space, the centre or the reference point of each bonding box is associated with three parameters (*x, y, z*), while in the image received from the camera, the original 3D space is reduced to two-dimensions of (*x, y*), and the depth parameter (*z*) is not available. In such a lowered-dimensional space, the direct use of the Euclidean distance criterion to measure inter-people distance estimation would be erroneous.

In order to apply a calibrated IPM transition, we first need to have a camera calibration by setting *z* = 0 to eliminate the perspective effect. We also need to know the camera location, its height, angle of view, as well as the optics specifications (i.e. the camera intrinsic parameters)^74^.

By applying the IMP, the 2D pixel points (*u, v*) will be mapped to the corresponding world coordinate points (*X_w_,Y_w_,Z_w_*):

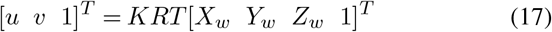

were *R* is the rotation matrix:

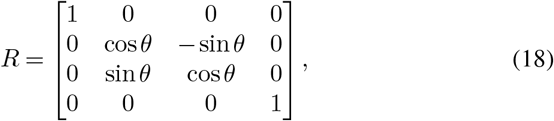

*T* is the translation matrix:

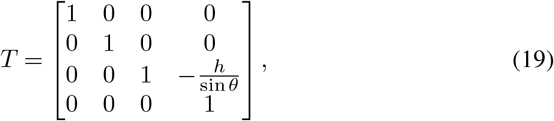

and *K*, the intrinsic parameters of the camera are shown by the following matrix:

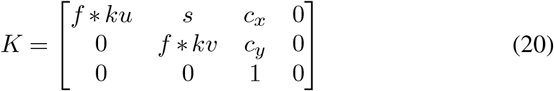

where *h* is the camera height, *f* is focal length, and *ku* and *kv* are the measured calibration coefficient values in horizontal and vertical pixel units, respectively. (*c_x_, c_y_*) is the principal point shifts that corrects the optical axis of the image plane.

The camera creates an image with a projection of threedimensional points in the world coordinate that falls on a retina plane. Using homogeneous coordinates, the relationship between three-dimensional points and the resulting image points of projection can be shown as follows:

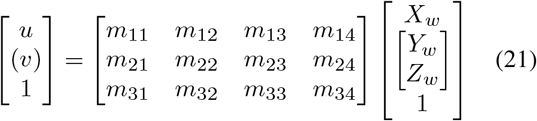

*were M ∊* ℝ^3×4^ is the transformation matrix with *m_ij_* elements in Equation 21, that maps the world coordinate points into the image points based on the camera location and the reference frame, provided by the Camera Intrinsic Matrix *K* (Eq. 20), Rotation Matrix *R* (Eq. 18) and the Translation Matrix *T* (Eq. 19).

Considering the camera image plane perpendicular to the *Z* access in the world coordinate system (i.e. *z* = 0) the dimensions of above equation can be reduced to the following form:

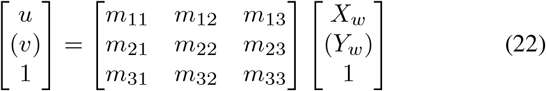

and finally transferring from the perspective space to inverse perspective space (BEV) can also be expressed in the following scalar form:

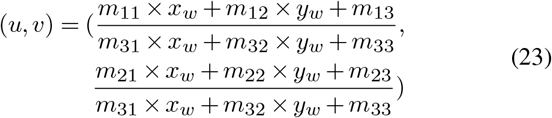

## 4 Experimental Results

In order to train the developed model, we considered a transfer learning approach by using pre-trained models on Microsoft COCO dataset^60^ followed by fine-tuning and optimisation of our YOLO-based model.

Four common multi-object annotated datasets including Pascal VOC^59^, COCO^60^, Image Net ILSVRC^76^, and Google Open Images^72^, were investigated in terms of the number of bounding boxes for human or person.

Figure 8 represents the sorted rank of object classes with the number of bounding boxes for each class in each dataset. In Google Open Images dataset (GOI) the class *Person* shows 4*^th^* rank, with nearly 10*^6^* annotated bounding boxes; richer than other three investigated datasets. In addition to the class *Person*, we also adopted four more classes of *Man, Woman, Boy, and Girl* from the GOI dataset for the human training purpose. This made a total number of 3,762,615 samples that we used from training, including 257,253 samples from the COCO dataset and 3,505,362 samples from the GOI dataset.

**Figure 8.**
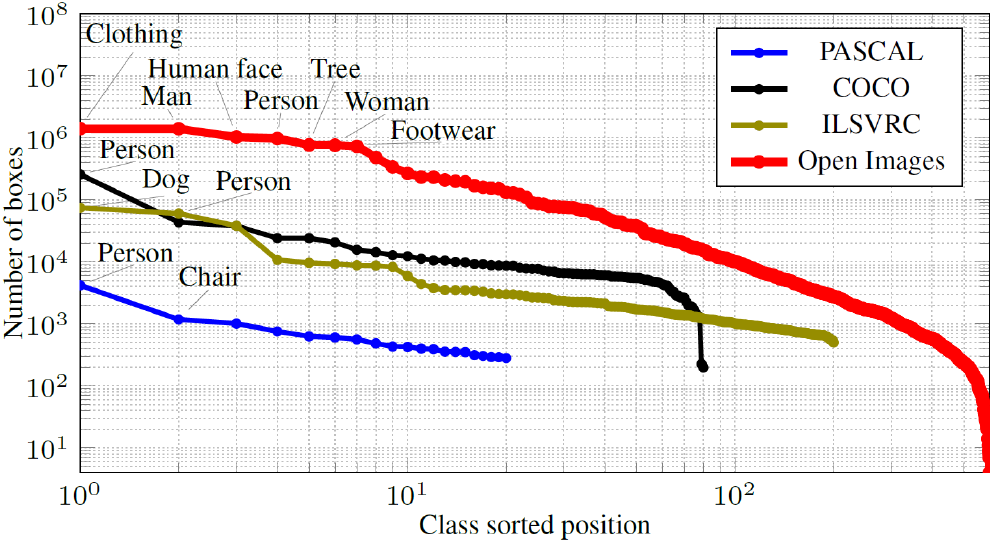
The number of annotated boxes per class in four common datasets. The horizontal axis is represented in logarithmic scale for better readability.

Figure 9 shows examples of annotated images from the Open Images Dataset. The figure illustrates the diversity of annotate persons including large and small bonding boxes, in far and near distances to the camera image plane, people occlusion, as well as variations in shades and lighting conditions.

**Figure 9.**
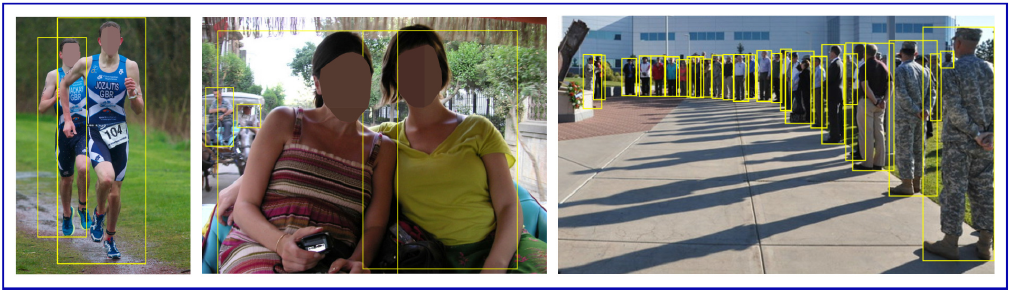
Examples of annotated images in Open Images dataset.

### 4.1 Performance Evaluation

In order to test the performance of the propose model, we used the Oxford Town Centre (OTC) dataset^29^ as a previously unseen and challenging dataset with very frequent cases of occlusions, overlaps, and crowded zones. The dataset also contains a good diversity of human specimens in terms of cloths and appearance in a real-world public place.

In order to provide a similar conditions for performance analysis of YOLO based models, we fine-tuned each model on human categories of the Google Open Images (GOI)^72^ data set. This has been done by removing the last layer of each model and placing a new layer (with random values of the uniform probability distribution) corresponding to a binary classification (presence or absence of a human). Furthermore, in order to provide an equal condition for the speed and generalizability, we also tested each of the trained models against the OTC dataset^29^.

We evaluated and compared our developed models against three common metrics of object detection in computer vision, including Precision Rate, Recall Rate, and FPS against three state-of-the art human/object detection methods.

All of the benchmarking tests and comparisons were conducted on the same hardware and software: a Windows 10based platform with an Intel^c^ Core ^TM^ i5-3570K Processor and an NVIDIA RTX 2080 GPU with CUDA version 10.1.

Figure 10 illustrates the development of loss function in training and validations phases for four versions of our DeepSOCIAL model with different backbone structures. The graphs confirm a fast yet smooth and stable transition for minimising the loss function in DS version after 1090 epochs where we reached to an optimal trade-off point for both the training and validation loss. Table 2 provides the details of each backbone and the outcome of the experimental results against three more state-of-the-art model on the OTC Dataset.

**Figure 10.**
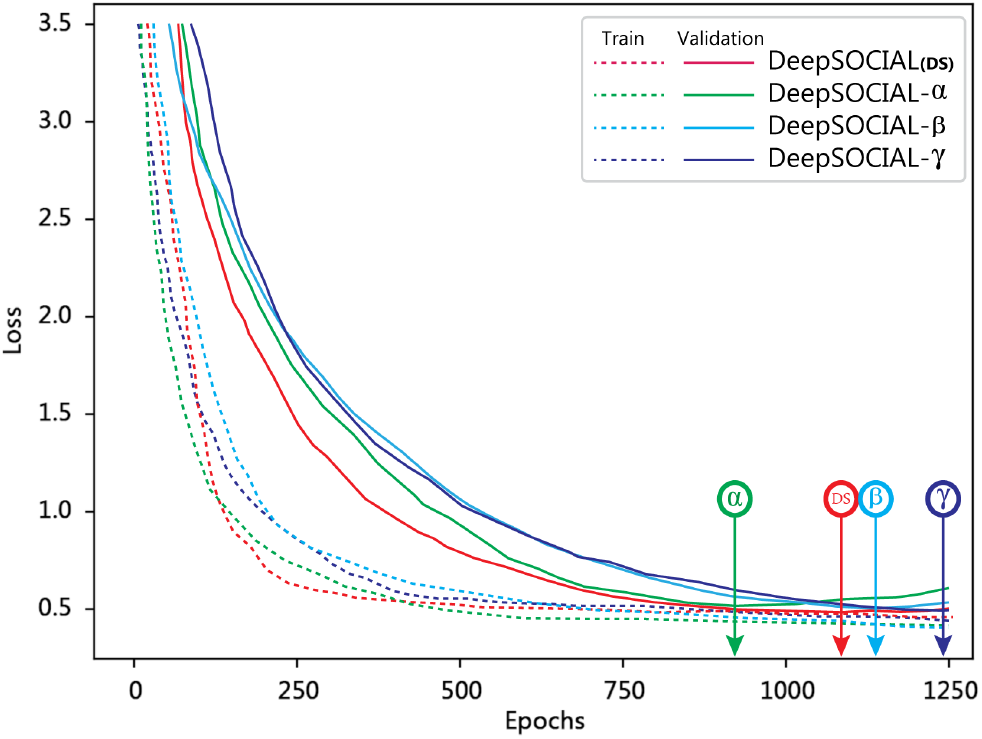
Training and Validation loss of the DeepSOCIAL models over the Open Images dataset.

**Table 2.** Speed, accuracy, and recall-rate comparison for seven DNN models on the Oxford Town Centre dataset.

| Method | Backbone | Precision | Recall | FPS |
| --- | --- | --- | --- | --- |
| DeepSOCIAL - $\alpha$ | CSPResNeXt50 - PANet - SPP - SAM | 99.8% | 96.7 | 23.8 |
| DeepSOCIAL - $\beta$ | CSPDarkNet53 - PANet - SPP | 99.5% | 97.1 | 24.1 |
| DeepSOCIAL - $\gamma$ | CSPResNeXt50 - PANet - SPP | 99.6% | 96.7 | 23.8 |
| **DeepSOCIAL (DS)** | CSPDarkNet53 - PANet - SPP - SAM | 99.8% | 97.6 | 24.1 |
| YOLOv3 | DarkNet53 | 84.6% | 68.2 | 23 |
| SSD | VGG-16 | 69.1% | 60.5 | 10 |
| Faster R-CNN | ResNet-50 | 96.9% | 83.0 | 3 |

Interestingly, the Faster-RCNN model shows good generalizability; however, its low speed is an issue which seems to be due to the computational cost of the “region proposal” technique. Since the system requires a real-time performance, any model with the speeds lower than 10 *fps* and/or a low level of accuracy may not be a suitable option for Social Distancing monitoring. Therefore, SSD and Faster-RCNN fail in this benchmarking assessment, despite their popularity in other applications. YOLOv3 and v4 provide relatively better results comparing the other models, and finally, the proposed DeepSOCIAL-DS model outperforms in terms of both speed and accuracy.

Figure 11 provides sample footage of the challenging scenarios when the people either enter or exit the scene, and only part of their body (e.g. their feet) are visible. The figure clearly indicates the strength of DeepSOCIAL in Row (a), comparing to the state-of-the-art. The bottom row (d) with blue bounding boxes shows the ground truth were some of the existing people with partial visibility are not annotated even in original ground truth dataset. Row (c), YOLOv3 shows a couple of more detections; however, the the IoU of the suggested bounding boxes are low and some of them can be counted as False Positives. Row (b), the standard YOLOv4-based detector, shows a significant improvement comparing to row (c) and is considered as the second-best. Row (a), the DeepSOCIAL, shows 10 more true positive detections (highlighted by vertical arrows) comparing to the seconds best approach.

**Figure 11.**
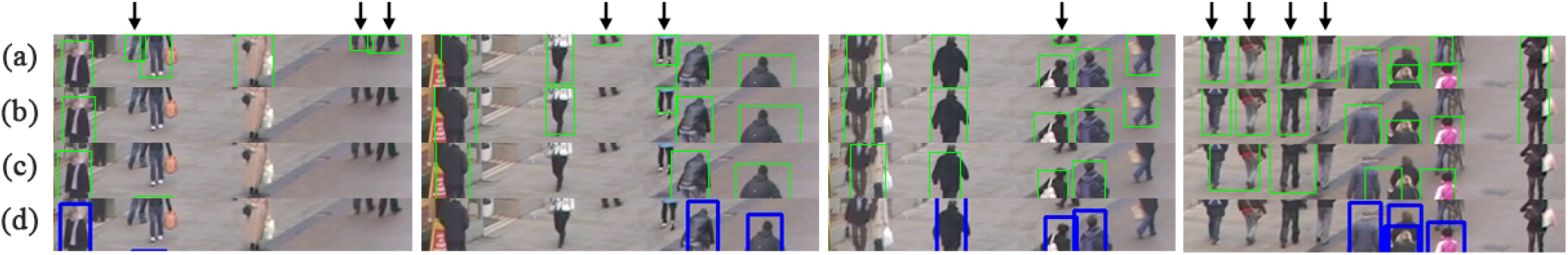
Human detection with partial visibility (missing upper body parts).

### 4.2 Social Distancing Evaluations

We considered the midpoint of the bottom edge of the detected bounding boxes as our reference points (i.e. shoes’ location). After the IPM, we would expect to have the location of each person, in the homogeneous space of BEV with a linear distance representation.

Any two people *P_i_, P_j_* with the Euclidean distance of smaller than *r* (i.e. the set restriction) in the BEV space are considered as contributors in social distancing violation:

Depending on the type of overlapping and the violation assessment criteria, we define a violation detection function *V* with the input parameters of a pixel metrics *£*, the set safe distance of *r* (e.g 6*ft* or ≈ 2*m*), the position of the query human *Hq*, and the closest surrounding person *Po*.

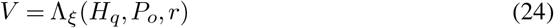

where ξ is number of pixels equal to 1.0 meter in the BEV space.

Figure 12.left (from Oxford Town Centre Dataset^29^) shows the detected peoples followed by the steps we have taken for inter-people distance estimation including tracking, IPM, homogeneous 360^ distance estimation, safe movements (people in green circles) and the violating people (with overlapping red circles):

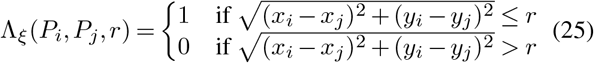

**Figure 12.**
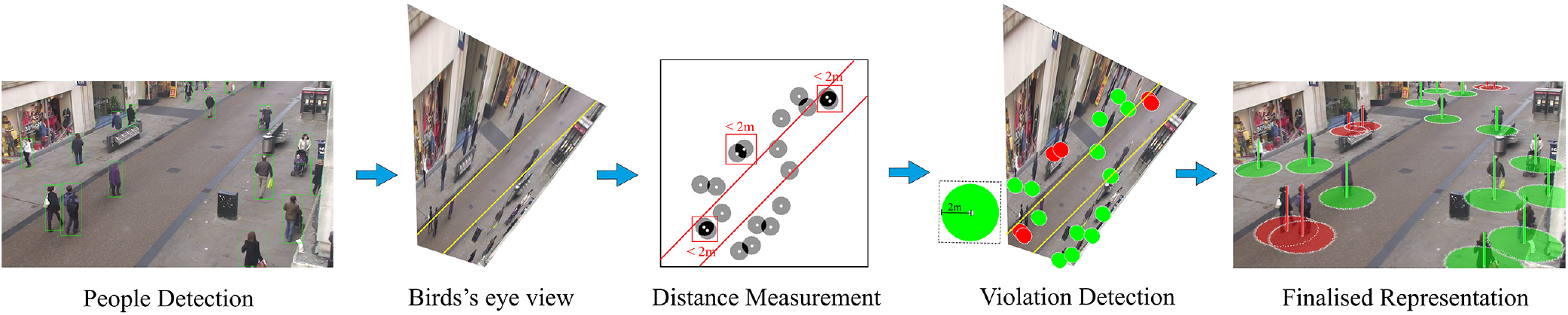
The summary of the steps taken in sections 3.1 to 3.3 for people detection, tracking, and distance estimation.

Regarding the Oxford Town Centre (OTC) dataset, every 10 pixels in the BEV space is equivalent to 98cm in the real world. Therefore, *r ≈* 2 × ξ and equal to 20 pixels. The inter-people distance measurement was measures based on the Euclidean *L*2 norm distance:

One of the controversial opinions that we received from health authorities were the way of dealing with family members and couples in social distancing monitoring. Some researchers believed social distancing should apply on every single individuals without any exceptions and others were advising the couples and family members can walk in a close proximity without being counted as breach of social distancing. In some countries such as in the UK and EU region the guideline allows two family members or a couple walk together without considering it as the breach of social distancing. We also considered a solution to activate the couple detection. This will be helpful when we aim at recognising risky zones based on the statistical analysis of overall movements and social distancing violations over a mid or long period (e.g. from few hours to few days).

Applying a temporal data analysis approach, we consider two individuals (*p_i_, p_j_*) as a couple, if they are less than *d* meters apart in an adjacency, for a *t_Λ_* of more than ε seconds. As an example, in Figure 13(a), we have identified people who have been less a meter apart from each other for more than *ε* = 5 seconds, in the same moving trajectory:

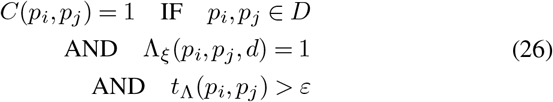

**Figure 13.**
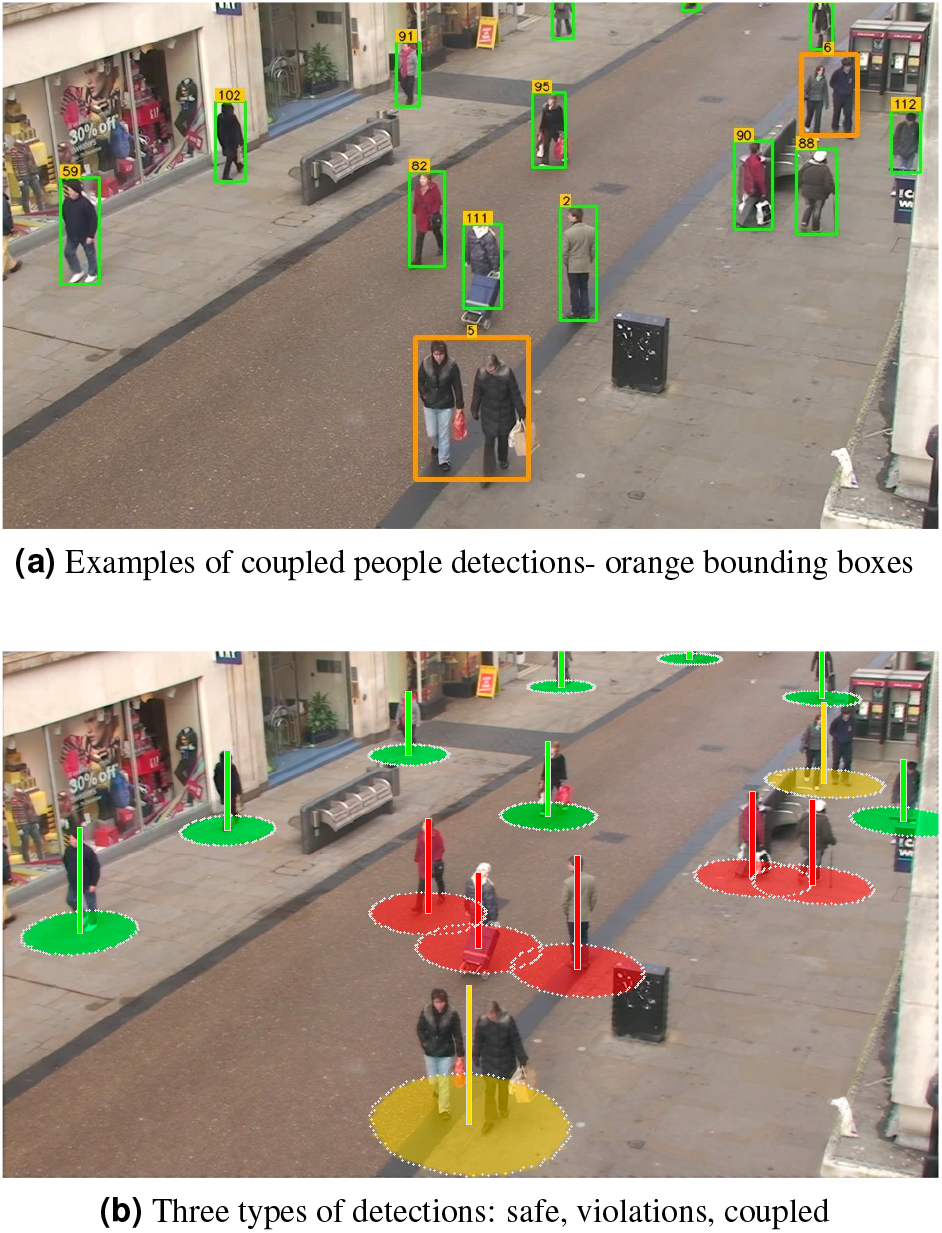
Social distancing violation detection for coupled people and individuals.

Figure 13(b) shows a sample of our representation for detected couples in a scene as well as multiple cases of social distancing violations. The yellow circles drawn for people diagnosed as couples have a radius of 3 meters to ensure a minimum safety distance of 2m for each of them to their left and right neighbours.

In cases were a breach occurs between two neighbouring couple, or between a couple and an individual, all of the involved people will turn to red status, regardless of being a couple or not.

The flexibility of our algorithm in considering different types of scenarios enables the policymakers and health authorities to proceed with different types of investigations and evaluations for the spread of the infection.

For example, Figure 14 from the Oxford Town Centre dataset, provides a basic statistics about the number of people in each frame, the number of people who do not observe the distancing, the number of social distancing violations without counting the coupled groups as violations.

**Figure 14.**
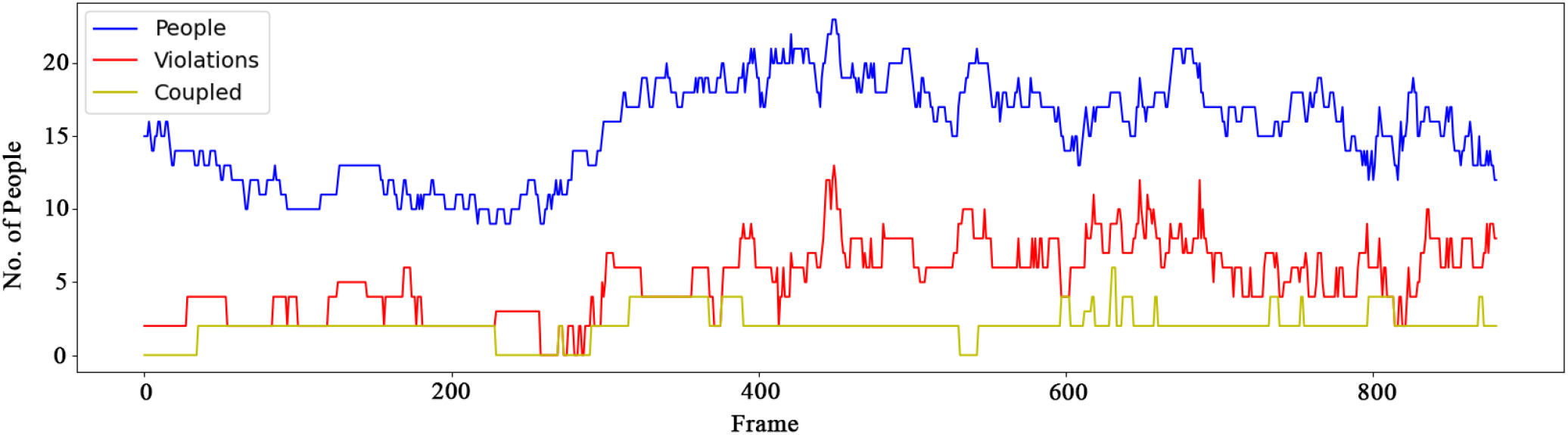
A 2D recording of the number of detected people in 900 frames from the OTC Dataset, as well as the number of violations and number of couples with no violations.

Regarding the coupled group we reached to an accuracy and recall rate of 98.7% and 23.9 *fps*, respectively which is

### 4.3 Zone-based Risk Assessment

We also tested the effectiveness of our model in assessing the long-term behaviour of the people. This can be valuable for health sector policy makers and governors to make timely decisions to save lives and reduce the consequent costs. Our experiments provided very interesting results that can be crucial to control the infection rates before it raises uncontrolled and unexpectedly.

In addition to people inter-distance measurement, we considered a long-term Spatio-temporal zone-based statistical analysis by tracking and logging the movement trajectory of people, density of each zone, total number of people who violated the social-distancing measures, the total time of the violations for each person and as the whole, identifying high-risk zones, and ultimately, creating an informative risk heat-map.

In order to perform the analysis, a 2-D grid matrix *G_t_* ∊ ℝ^*w* ×^ *^h^* (initially filled by zero) is created to keep the latest location of individuals using the input image sequences. *G_t_* represents the status of the matrix at time *t* and ω and *h* are the width and height of the input image *I*, respectively.

To consider environmental noise and better visualisation of the intended heat-map, every person was associated with a 3 × 3 Gaussian kernel *k*:

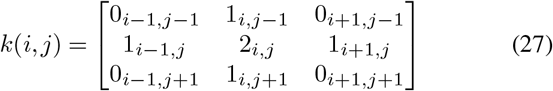

were *i* and *j* indicate the centre point of the predicted box for each individual, *p*.

The grid matrix *G* will be updated for every new frame and accumulates the latest information of the detected people.

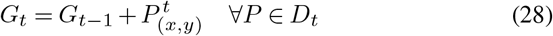

Figure 15 shows a sample representation of the accumulated tracking map after 500 frames of continuous people detection.

**Figure 15.**
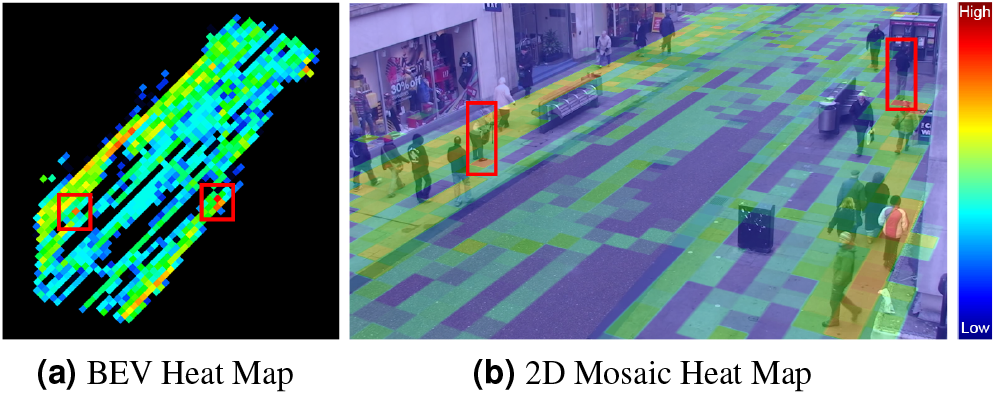
Accumulated tracking maps after 500 frames.

Since COVID-19 is an airborne virus, breathing by any static person in a fixed location can increase the density of the contamination on that point (assuming the person may carry the COVID-19), particularly in covered places with minimal ventilation. Therefore we can assign a more contamination weight to the steady state people.

Figure 15(b) and 15(a) shows two instances of cases where two people have been steady in two particular location of the grid for a long period; hence, the heat map is turning to red for those locations. Both sidewalks also show stronger heatmap than the middle of the street due to the higher traffic of people movements. In general, the more red grids potentially indicates more risky spots.

In addition to people raw movement and tracking data, that would be more beneficial analyse the people who particularly violated the social distancing measures.

In order to have a comprehensive set of information we aim to represent a long term heat-map of the environment based on the combination of accumulated detections, movements, steady states people, and total number of breaches of the social-distancing. This helps to identify risky zones, or redesign the layout of the environment to make it a safer place, or to apply more restrictions and/or limited access to particular zones. The restriction rules may vary depending on the application and the nature of the environment (e.g. this can vary in a hospital compared to a school).

Applying the social distancing violation criteria as per Eq. 24, we identify each individual in one of the following categories:

- **Safe:** All people who observe the social distancing (green circles).

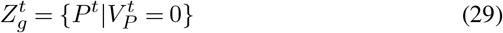
- **High-risk:** All people who violate the social distancing (red circles).

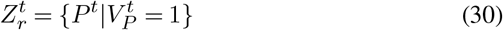
- **Potentially risky:** Those people who move together (yellow circles) and are identified as coupled. Any two people in a coupled group are considered as one identity as long as they do not breach the social distancing measures with their neighbouring people. The social distancing radius for coupled people is set as 3 meters.

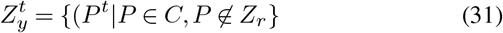

We also use a 3-D violation matrix *S ∊* ℝ^*w* × *h* × 3^ to record the location of breaches for each person and its type (red or yellow):

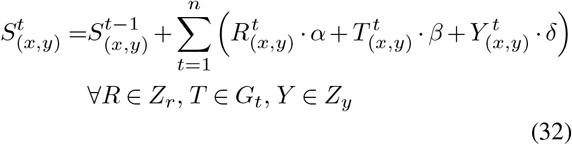

were *R*, *T*, and *Y* indicates cases with a violation, tracked people, and couples, respectively. *α, β*, and δ are the relative coefficients that can be set by health-related researches depending on the importance of each factor in spreading the virus.

In order to visualise the 3D heat map of safe and red zones, we normalise the collected data in Eq. 32 as follows:

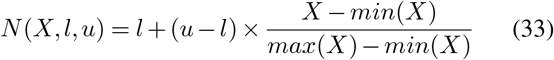

where X is the non-normalised values, *l* and *u* are the lower bound and upper bound of the normalisation matrix that we use to define the Hue colour range on the HSV channel.

Figure 16 shows a visualised output of the discussed experiments including tracking, 2-D and 3-D analytical heat-map of risky zones for the Oxford Town Centre dataset in HSV colour space:

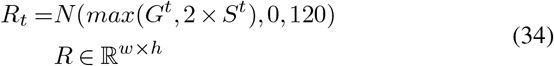

**Figure 16.**
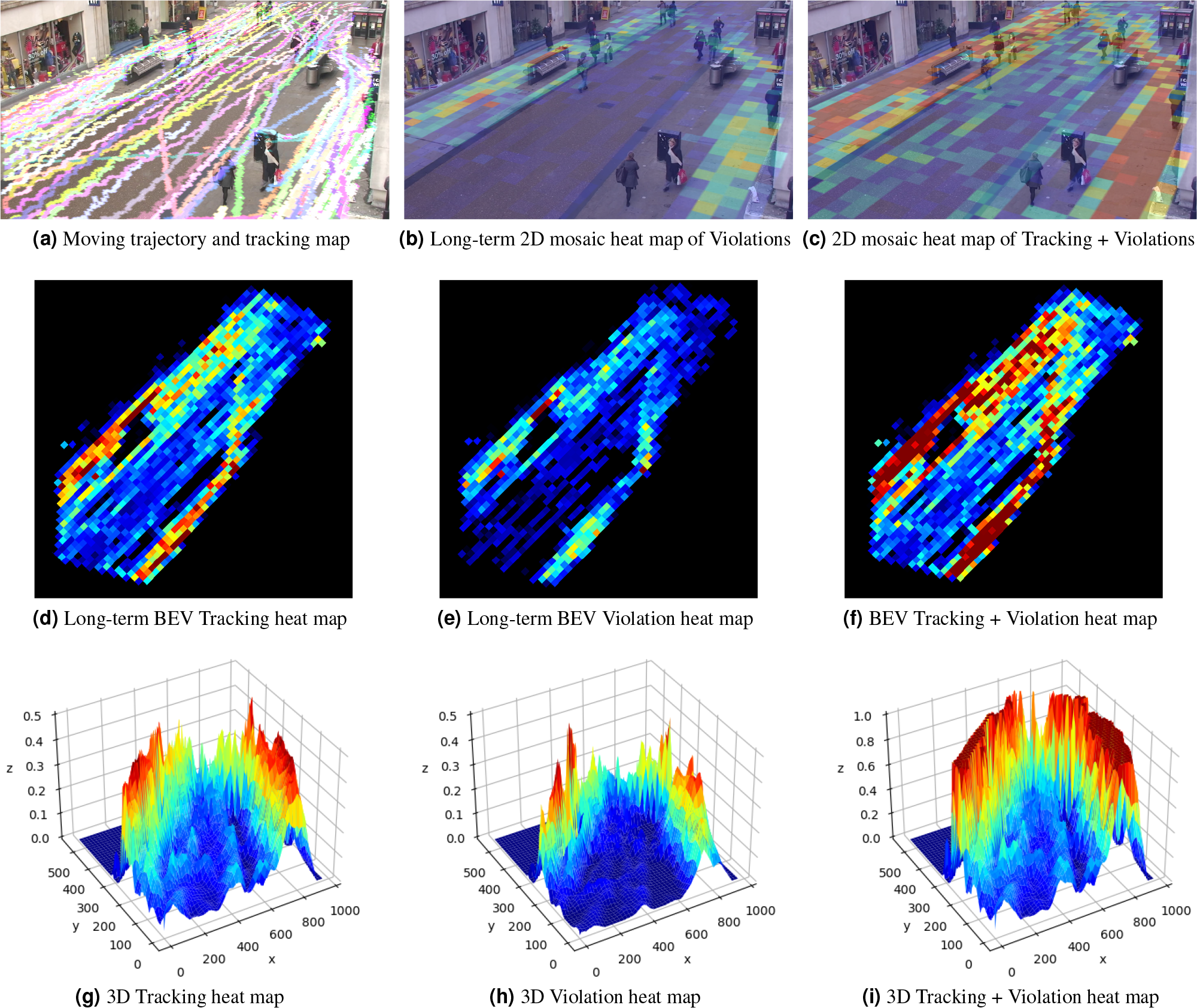
Data analysis based on people detections, tracking, movements, and breaches of social distancing measurements.

Figure 16(a) shows the tracking paths of the passed people after 2500 frames. Figure 16(b) illustrate a mosaic heatmap of people with social distancing violations only. Figure 16(c) shows the summed heatmap of social distancing violations and tracking. Figures 16(d), (e), and (f) show the birds-eye view heatmap of long term-tracking, violations, and the mixed heatmap, respectively. Figures 16(g), (h), and (i) are the corresponding 3D representation of the same heatmaps in the second row, for better visualisation of safe and risky zones.

The above configuration was an accumulating approach were all the violations and risky behaviours were added together in order to highlight potentially risky zones in a covered area with poor ventilation.

We also thought about cases when there exist a good chance of ventilation were the spread of virus would not be necessarily accumulative. In such cases we considered both increasing and decreasing counts depending on the overall time spent by each individual in each grid cell of the image, as well as the total absence time of individuals which potentially allows bring down the level of contamination.

Figure 17 which we named it as crowd map shows the 2D and 3D representation of violations and risk heat maps where we applied both increasing and decreasing contamination trends. The first row of the image represents a single frame analysis with two peak zones. As can be seen, those zones belong to two crowded zones were two large groups of people are walking together and breach the social distancing rule. However, in other parts of the street a minimal level or risk is identified. This is due to large inter people distances and a consequent time gaps which allows breathing and therefore a decreasing rate of contamination.

**Figure 17.**
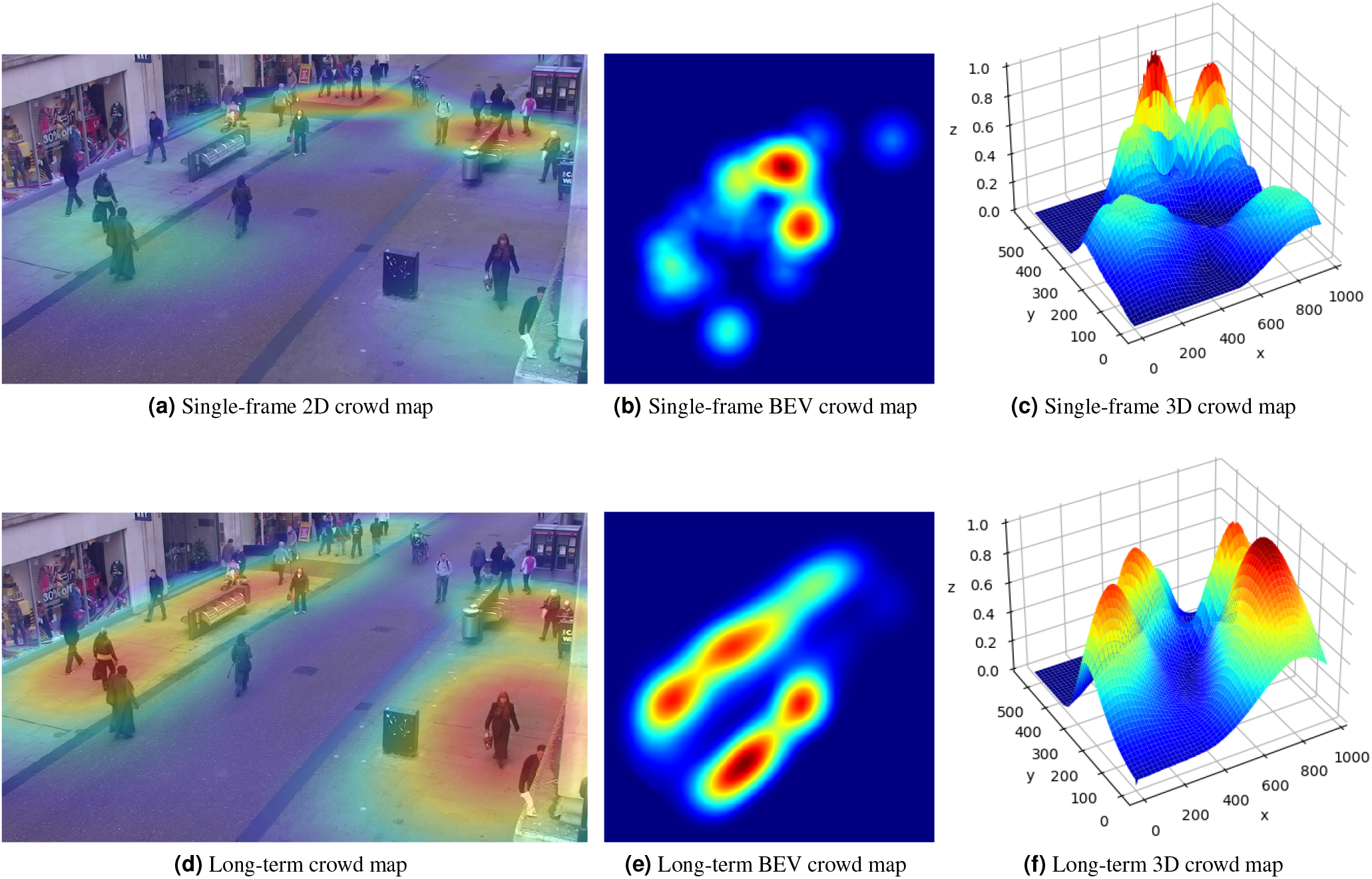
Single frame vs. Long-term crowd map (2D, BEV, 3D).

The second row of Figure 17 shows a long-term crow map which does not necessarily depend on the current frame. This can be a weighted averaging over all of the previous singleframe crow maps.

One of the extra research questions in Figure 16 and 17 is how to define appropriate averaging weights and coefficients (α*, β, γ)* in Equation 32 and how to normalise the maps over the time. This is out of the scope of this research and needs further studies. However, here we aimed at showing the feasibility of considering a diversity of cases using the proposed method with a high level of confidence and accuracy in social distancing monitoring and risk assessment with the help of AI and Computer Vision.

## 5 Conclusion

We proposed a Deep Neural Network-Based human detector model called DeepSOCIAL to detect and track static and dynamic people in public places in order to monitor social distancing metrics in COVID-19 era and beyond. We utilised a CSPDarkNet53 backbone along with an SPP/PAN and SAM neck, Mish activation function, and Complete IoU loss function and developed an efficient and accurate human detector, applicable in various environments using any type of CCTV surveillance cameras. The system was able to perform in a variety of challenges including, occlusion, lighting variations, shades, and partial visibility. The proposed method was evaluated using large and comprehensive datasets and proved a major development in terms of accuracy and speed compared to three state-of-the-art techniques. The system performed real-time using a basic hardware and GPU platform.

We also conducted a zone based infection risk assessment and analysis to the benefit of the health authorities and governments. The outcome of this research is applicable for a wider community of researchers not only in computer vision, AI, and health sectors, but also in other industrial applications such as pedestrian detection in driver assistance systems, autonomous vehicles, anomaly behaviour detections, and variety of surveillance security systems.

## Data Availability

All data will be available upon approval

